# Systematic Review and Meta-Analysis: Relationships Between Attention-Deficit/Hyperactivity Disorder and Daytime Urinary Symptoms in Children

**DOI:** 10.1101/2020.08.28.20183541

**Authors:** Behrang Mahjani, Lotta Renström Koskela, Christina Gustavsson Mahjani, Magdalena Janecka, Anita Batuure, Christina M. Hultman, Abraham Reichenberg, Joseph D. Buxbaum, Olof Akre, Dorothy E. Grice

## Abstract

Lower urinary tract symptoms (LUTS), e.g., urinary frequency, pressure, urgency, and overactive bladder syndrome, are commonly reported in children with attention-deficit/hyperactivity disorder (ADHD). Understanding the co-occurrence of these conditions has implications regarding clinical approaches, treatments, and improved quality of life. We conducted a systematic review and meta-analysis to examine the relationships between LUTS and ADHD in children.

We searched for articles published between January 1990 and July 2019, in PubMed, CENTRAL, and PsycNet. Two authors independently screened all articles and extracted data. We performed random-effect meta-analyses for ADHD with pooled outcomes for LUTS.

We identified 117 relevant articles in the literature and 17 articles fulfilled the inclusion criteria for the systematic review, of which, 5 articles had sufficient data for meta-analysis. Examining ADHD among individuals with LUTS, the odds ratio was 2.99 (95% CI: 1.13,7.88, P < 0.001), compared to controls. In multiple studies, the mean overall score for LUTS, using a standardized measure, was significantly higher in patients with ADHD in comparison to controls, and the severity of ADHD was positively associated with the severity of LUTS. Younger age in children was correlated with a higher LUTS score. Different subtypes of urinary incontinence demonstrated differences in behavioral problems and psychiatric comorbidity. Sex differences in LUTS were not consistent across articles.

Our results indicate clinically significant associations between ADHD and LUTS in children. Because LUTS and ADHD are common disorders in children, clinicians should be aware of these associations as they inform optimal assessment and treatment strategies.

## 1. Introduction

Attention-deficit/hyperactivity disorder (ADHD) is one of the most commonly diagnosed psychiatric disorders in children [1]. Approximately 5-9.4% of children are diagnosed with ADHD, with a higher prevalence of the disorder in males than females [1, 2]. The core diagnosis requires ongoing patterns of behavior that represent inattention and/or hyperactivity-impulsivity and interfere with development or expected functioning [3]. However, ADHD is often comorbid with other psychiatric disorders [4, 5] and medical conditions. Comorbidity with lower urinary tract symptoms [6, 7] and overactive bladder syndrome [8, 9] has been reported.

Lower urinary tract symptoms (LUTS) is a term that covers symptoms (e.g., storage, voiding, post-micturition, pain) that result from conditions affecting the bladder and urethra. Storage symptoms manifest as urinary urgency, frequency, nocturia, or incontinence. Incontinence can be divided into daytime incontinence and nighttime incontinence (i.e., enuresis). Daytime incontinence includes urge incontinence, stress incontinence, and mixed incontinence [10]. The population frequency of LUTS in children is between 9-40%, and varies due to definitions and subcategories of LUTS [11, 12]. Overactive bladder syndrome (OAB) is a symptom-based clinical diagnosis and is characterized by excessive urgency and frequency of urination. OAB has a prevalence of 5-12% in children, and at least a third of affected children continue into adulthood with similar complaints [13]. LUTS and OAB have significant adverse effects on the overall quality of life. They can affect children’s social and emotional development and increase risk for social isolation and teasing.

To date, no meta-analyses of associations between ADHD and LUTS have been published. We hypothesize that there is a clinically significant association between ADHD and lower urinary tract symptoms. Understanding the association between these two disorders is expected to have a positive impact on patient quality of life by providing the foundation for early preventive interventions, which is currently lacking, and more nuanced treatments. Therefore, we conducted a systematic review and meta-analysis to explore this association.

## 2. Methods

### 2.1 Search strategy and selection criteria

We followed the PRISMA protocol [14] and registered our approach in PROSPERO [15] (ID= CRD42019118607). Figure 1 illustrates our search strategy to find the qualifying articles, which includes identification, screening, and eligibility. *Identification* We identified all articles with results in humans that included our search terms referencing ADHD or LUTS in the title, and ADHD and LUTS in the title or abstract; *Screening* At least two of the authors (BM, MJ, AB) independently screened all articles based on titles and abstracts (see below for the exclusion criteria). Twenty articles were randomly chosen and re-screened by a different author (DG), as a quality control measure; *Eligibility* At least two of the authors (CM, AB, BM) independently reviewed the full text of the articles that passed screening. Excluded articles and reviewer conflicts were assessed independently by a different author. Twenty articles were randomly chosen and re-screened by a different author (DG). We used DistillerSR (Evidence Partners, Ottawa, Canada) software for screening the articles. For LUTS outcomes, we included overactive bladder, bladder pain syndrome, and daytime incontinence. We included all diagnoses and symptoms, even where the specific reference to ICD/DSM was missing, provided a standardized measure was used in the study. We systematically searched for articles in PubMed, CENTRAL, PsycNet, and Google Scholar. Specific search terms used for PubMed search were: ((urinary) OR (lower urinary tract) OR (overactive bladder) OR (bladder pain syndrome) OR (incontinence)) AND (attention deficit hyperactivity) AND (urinary[Title] OR lower urinary tract[Title] OR overactive bladder[Title] OR bladder pain syndrome[Title] OR incontinence[Title] OR attention deficit hyperactivity[Title]) NOT rats[Title] NOT rat[Title] NOT dog[Title] NOT dogs[Title] NOT rabbits[Title] NOT rabbit[Title] NOT mice[Title]. We used Google Scholar to ascertain if any articles were missed in the PubMed, CENTRAL and PsycNet searches. We performed 2 searches in Google Scholar using different keywords (details in Supplement 2). For each search, we screened the first 100 articles to identify articles that were not found previously. We also examined all references from relevant review articles.

**Figure 1.**
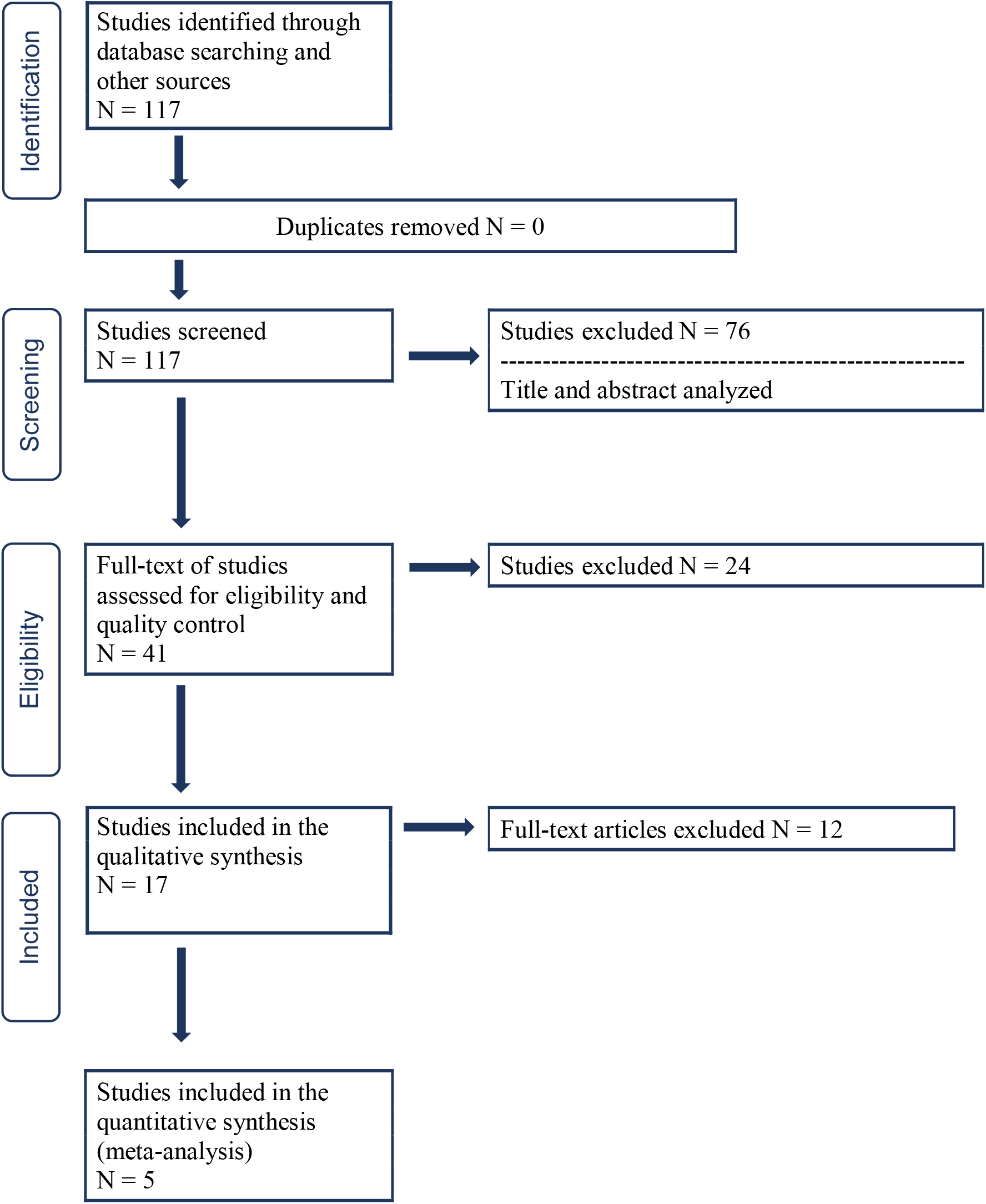
PRISMA details

We considered peer-reviewed, published studies (including studies published online ahead of print) in English (or translated into English) from January 1, 1990, to July 15, 2019. We excluded all studies published as editorials or commentaries. We excluded treatment trials, studies with less than 20 participants, studies where all participants were ≥ 50 years old (due to the impact of aging on urinary symptoms), studies reporting only nocturnal enuresis, and studies in which it was not possible to separate the results for daytime incontinence from those for nocturnal enuresis/nighttime incontinence. We excluded LUTS associated with physical trauma, cancer, sexually-transmitted infection, congenital malformation of the urinary tract, post-operative disorders, those related primarily to pregnancy, dysfunction secondary to aging, and medications that promote urination/water retention. We excluded outcomes associated with kidney malfunction. Observational studies were included in the review (cohort studies, case-control studies, and cross-sectional studies). We did not include ecological, treatment, or case series studies.

### 2.2 Data analysis

For each study, we extracted the number of exposed and unexposed individuals with LUTS and ADHD to calculate the odds ratio. We excluded articles that did not provide the necessary data for meta-analysis. If multiple studies analyzed the same data, then we chose only one study. We performed meta-analyses using random-effects models and evaluated publication bias using funnel plots and applied Egger’s test [16]. We used *metafor* package in R for the meta-analysis [17].

## 3. Results

We identified 117 articles published from January 1, 1990 to July 15, 2019 that met the inclusion criteria (Figure 2, *Identification*). After the review of titles, abstracts, and full texts, 17 articles were included in the qualitative and quantitative synthesis (Table 1). Of those articles, 5 had the required information for meta-analysis (Table 1). Urinary storage symptoms (urinary urgency, urinary frequency, daytime urinary incontinence (DUI)) and voiding problems were the most commonly reported LUTS that co-occur with psychiatric disorders.

**Figure 2.**
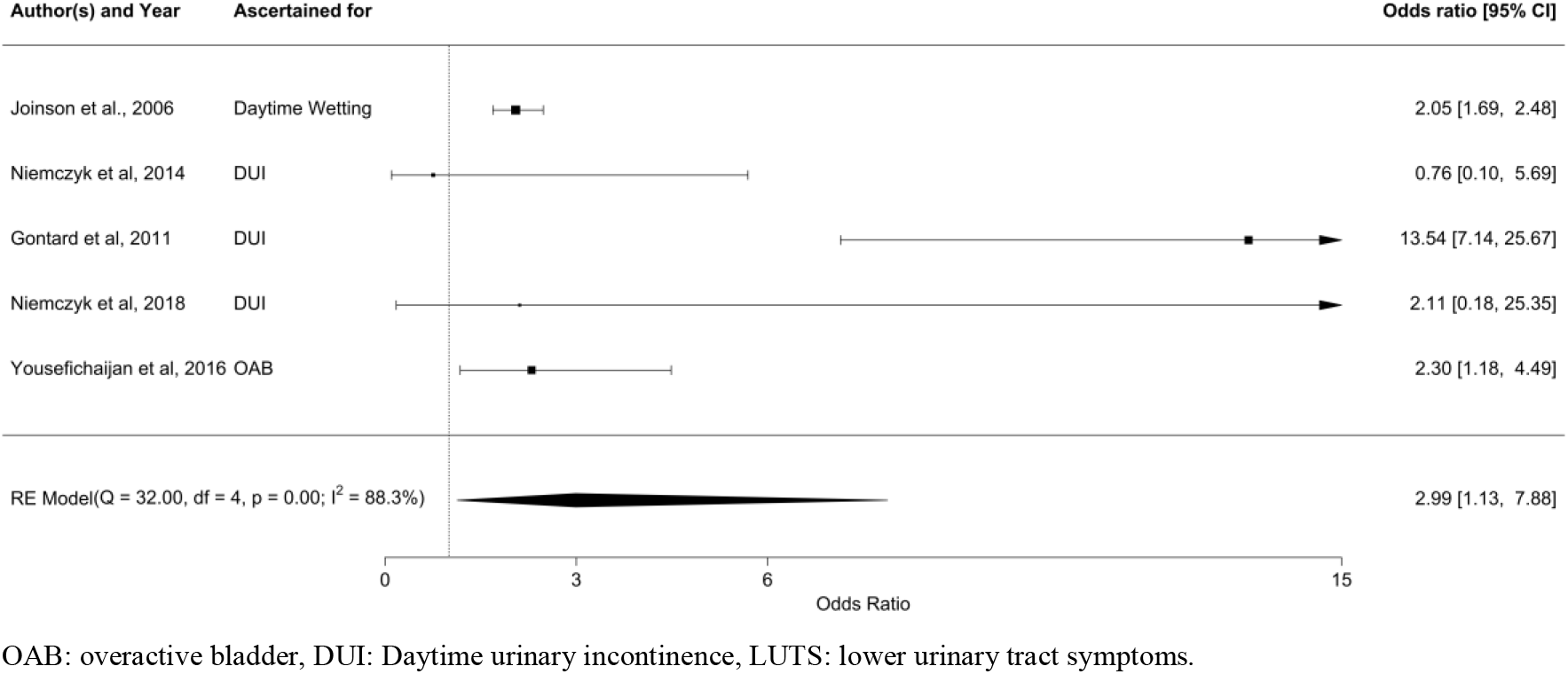
Forest plot for ADHD individuals with LUTS.

**Table 1.** Characteristics of included studies (*n* = 17)

| Study | LUTS Assessment (scale) | ADHD Assessment Methods | Age | Notes | Study design |
| --- | --- | --- | --- | --- | --- |
| Burgu et al. [6] | LUTSS<sup>1</sup> | CPRS-R<sup>2</sup> | 6-17 | Age matched controls | Case-control |
| Duel et al. [7] | DVSS<sup>3</sup> | Clinical diagnosis at Child Development Center | 6-12 | Controls from general pediatrics clinic | Case-control |
| Gontard et al. [19] | Structured interview | CBCL<sup>4</sup> and clinical diagnosis | Mean: 6.22 |  | Cohort study, county-based population |
| Gor et al. [33] | Clinical examination for daytime voiding symptoms | Clinical diagnosis | Mean: 8.6 |  | Retrospective observational study |
| Joinson et al. [20] | Parent report | DAWBA<sup>5</sup> | 7-9 | Parents completed a postal questionnaire | Population-based longitudinal study |
| Niemczyk et. al [29] | Parent report | Parent report | 4-8 |  | Cross-sectional study |
| Niemczyk et al. [32] | Clinical incontinence examination | CBCL<sup>4</sup> | 5-13 | Emotion processing in children with DUI | Case-control |
| Oliver et al. [21] | LUTSS<sup>1</sup> | Parent report | 6-17 |  | Cross-sectional study |
| Özen et al. [8] | VDSS<sup>6</sup> | Clinical diagnosis by child psychiatrist | 6-18 | Case only | Retrospective observational study |
| Schast et al. [22] | PLUSS<sup>7</sup> | Parent report | 5-17 |  | Cross-sectional study |
| Sureshkumar et al. [23] | Parent report (validated) | Parent report (validated) | Mean age: 7.3 |  | Cross-sectional study |
| Kuizenga-Wessel et al. [35] | Standardized questionnaires including urinary incontinence | CBCL<sup>4</sup> and VvGK<sup>8</sup> | Median: 9.5 |  | Cross-sectional cohort study |
| Wolfe-Christensen et al. [24] | DVSS<sup>3</sup> | PSC<sup>9</sup> and clinical diagnosis by psychologist | 4-16 | Retrospective chart review | Cross-sectional study |
| Yang et al. [25] | DVSS<sup>3</sup>, Chinese version | SNAP-IV<sup>10</sup>, Chinese version | 4-14 | Case only | Cross-sectional study |
| Yousefichaijan et al. [9] | Clinical presentation, imaging, and exclusion of other causes of voiding dysfunction, led to overactive bladder diagnosis | CPRS-R and DSM IV-TR criteria | Mean 8.12 |  | Case-control |
| Zink et al. [26] | Referred to a specialized outpatient clinic for elimination disorders | CBCL<sup>4</sup> and ICD-10 | 5-16 |  | Cross-sectional study |
| Sureshkumar et al. [27] | Parent report (validated) | Parent report (validated) | Mean 7.3 |  | Cross-sectional study |

<sup>1</sup>LUTSS: Lower Urinary Tract Symptom Score; CPRS-R<sup>2</sup>: Conners's Parent Rating Scale; <sup>3</sup>DVSS: Dysfunctional Voiding Symptom Survey;
<sup>4</sup>CBCL: Child Behavior Checklist; <sup>5</sup>DAWBA: Development and Well-Being Assessment; <sup>6</sup>VDSS: Voiding Dysfunction Symptom Score; <sup>7</sup>PLUSS: Pediatric Lower Urinary Symptom Score; <sup>8</sup>VvGK: (Dutch questionnaire based on the American Disruptive Behavior Disorder rating scale); <sup>9</sup>PSC: Pediatric Symptom Checklist; SNAP-IV<sup>10</sup>: Swanson, Nolan and Pelham-IV Teacher and Parent Rating Scale

### 3.1 Qualitative synthesis

For the 17 articles eligible for qualitative synthesis, several reported that patients with ADHD have a higher incidence of LUTS compared to age-and sex-matched controls [6, 7]. In a study of 62 children (6-17 age range) with ADHD and 124 healthy controls, the mean total LUTS score (measured using the Urinary Tract Symptom Score, LUTTS) was 11.1 ± 2.9 in patients with ADHD and 3.2 ± 1.3 in controls (p < 0.001) [6]. LUTTS contains 14 questions, with the total score ranged between 0 and 56 [18]. Similar findings emerge from a study of 28 cases (23 boys and 5 girls; ages 6-12 years) and 22 controls using a modified Dysfunctional Voiding Symptom Scale (DVSS). The modified DVSS was designed to quantify voiding symptoms in children using 10 questions, scored on a 0 to 4 scale. In this study, the mean overall score for LUTS using the modified DVSS in boys with ADHD and controls was 14.83 ± 3.68 and 6.00 ± 5.74 (p < 0.0005), respectively. Overall, children of both sexes with ADHD had statistically significant higher overall DVSS scores [7].

Multiple studies reported that in patients with LUTS there was a higher incidence of ADHD compared to matched controls [8, 9, 27, 19–26]. Joinson et al. conducted one of the largest population-based analyses of the psychological problems associated with daytime incontinence in children using the Avon Longitudinal Study of Parents and Children (ALSPAC) cohort [20]. ALSPAC is a large population-based longitudinal study of children born in the former county of Avon, England, during 1991 and 1992 [28]. A self-report questionnaire asked parents a set of questions about their child’s toileting behavior. The Development and Well-Being Assessment (DAWBA) was also included in the questionnaire [28]. Among 8213 children (7-9 age range), 643 children (7.8%) had daytime wetting. Children with daytime wetting also had a higher rate for each of the parent-reported psychological problems compared to children with no daytime wetting. The largest difference between those with daytime wetting and those without was in attention and activity problems (24.8%).

Two large studies in Germany similarly reported a higher rate of ADHD among children with LUTS. In a school entry medical examination of 2,348 children in Saarbrücken county, parents completed a 32-item questionnaire regarding incontinence, oppositional defiant disorder, and ADHD symptoms [29]. Boys and girls had similar rates of daytime incontinence, and 10.3% of children with incontinence had ADHD. In another school entry medical examination of 1,391 children in the Saarpfalz Kreis county, ADHD symptoms (using the Child Behavior Checklist [30]) were more common in children with urinary incontinence than non-wetting children (16.8% vs. 3.4%).

Additional support for the positive association between ADHD and LUTS comes from a large study in Sydney, Australia. In this population-based study of 2,856 children, urinary frequency and daytime incontinence were grouped into very mild, mild, moderate, and severe categories. Severe daytime urinary incontinence was positively associated with ADHD with an odds ratio of 4.8 (95%CI, 2-11.9) [23].

In a urology clinic sample (n=120), Yang et al. examined hyperactivity in children with untreated LUTS [25]. Boys with higher scores of ADHD hyperactive subtype had a higher DVSS score [31].

Multiple studies have analyzed a range of behavioral comorbidities, including ADHD, in children with LUTS who present at urology clinics. Of 600 children presenting to an outpatient urology clinic, 15.2% met the clinical cut-off for significant psychosocial difficulties utilizing parent report the Pediatric Symptom Checklist. ADHD was the most commonly reported disorder (14%), followed by depression and anxiety [24]. In another study of 358 children (6 to 17 years old) with non-neurogenic lower urinary tract dysfunction, younger age was correlated with a higher LUTS score (r=-0.34, p < 0.0001, measured using LUTTS) [21] and ADHD was the most prevalent psychiatric disorder (8.4%). Schast et al. studied 351 voiding clinic patients at the Children’s Hospital of Philadelphia, USA, and found that 25% were diagnosed with a mental or behavioral health problem, and, ADHD was the most commonly reported diagnosis (13%) [22] a higher rate than a recent US national survey that reported 9.4% ADHD prevalence [2]. In a study of 166 children referred to a specialized outpatient clinic, different subtypes of urinary incontinence reported different behavioral problems and psychiatric comorbidity; children with voiding postponement had the highest rates of psychiatric comorbidities, including ADHD [26].

Özen et al. studied the association between lower urinary tract dysfunction and psychiatric disorders in 156 children and highlights the importance of screening tests for psychological problems among children with LUTS. Among children with LUTS, ADHD was the most common psychiatric disorder (16.1%). This study is one of the very few studies that include a subsample of individuals with OAB (76 children). 19.7% of children with OAB had ADHD [8]. In another study of OAB, Yousefichaijan et al. investigated the risk of ADHD in 92 children with OAB and 92 healthy controls [9]. The prevalence of ADHD among the cases was significantly higher than the controls (35.9% vs. 19.6%; P = 0.021).

### 3.2 Meta-analysis

Our review yielded 5 articles that qualified for meta-analysis. The characteristics of these studies are present in Table 1. The odds ratio using random effect models for ADHD among individuals with LUTS was 2.99 (95% CI: 1.13,7.88, P < 0.001; Figure 1).

The large value of Cochran’s Q test, I^2^ index, and visual inspection of the funnel plot (Figure 2 and Figure S1) suggest a high heterogeneity between studies. The heterogeneity may be related to bias from publication, reporting, and selection, as supported by two studies plotting outside the funnel outline in the funnel plot.

## 4. Discussion

This systematic review and meta-analysis demonstrate a strong association between ADHD and LUTS. The odds ratio for ADHD amongst children with a LUTS diagnosis was 2.99 (95% CI: 1.13,7.88, P < 0.001). Different subtypes of urinary incontinence demonstrated differences in behavioral problems and psychiatric comorbidity; however, multiple studies indicate that ADHD is the most common psychiatric disorder among children with LUTS and occurs at rates higher than expected based on population prevalence. Although there were insufficient data for meta-analysis, multiple studies reported that the mean score for LUTS (measured via the LUTTS and DVSS) was significantly higher in patients with ADHD in comparison to controls, and the severity of ADHD was positively associated with the severity of LUTS. In addition, younger age in children was positively correlated with a higher LUTS score. We did not observe a consistent pattern for sex differences in LUTS across article reviews.

The present meta-analyses had several strengths and some limitations. This study is the first systematic review and meta-analysis analyzing the association between ADHD and LUTS in children. We included multiple case-control and cohort studies with a relatively large sample size and low risk of bias. However, only 5 studies were included in the meta-analysis; therefore, the potential effect of publication bias and small study effects should be considered when interpreting the results. Comparison of Cochran’s Q test and I^2^ index suggested a high degree of heterogeneity in the studies. We believe that the leading cause of heterogeneity was the variation in the definition of LUTS and ADHD. Diagnostic criteria and definitions of LUTS and ADHD were not always consistent or clear across the studies. Some studies used a clinical diagnosis, while others measured LUTS and ADHD symptoms using standardized assessments. While the Egger test was not statistically significant (p-value=0.50), there were small numbers of studies, that reduced its power. With visual inspection of the funnel plot, it appears that smaller studies without statistically significant effects may not have been published.

In summary, our results indicate clinically significant associations between ADHD and LUTS in children. LUTS is a common disorder in children, and clinicians should be aware of this association because this can interfere with treatment or inform optimal treatments. In addition, the presence of ADHD may impact performance of behavioral therapies for LUTS. Our analyses also highlight a significant gap in the literature. Studies of LUTS in children ascertained for ADHD were rare. Additionally, there were no studies using national registers, in which there is potentially less ascertainment and confounding bias. Despite the significant associations between forms of LUTS and psychiatric disorders, the factors underlying the significant co-occurrence of these conditions remain mostly unidentified. We also know little about the direction of risk and impact on treatment. Undoubtedly, optimal treatment for LUTS needs to incorporate the psychiatric dimension, requiring a more precise understanding of these relationships. Understanding the causes of the observed relationships between ADHD and LUTS, biological or behavioral, is an important topic for future study.

## Data Availability

data available upon request

## Availability of data and material

Data are available in the literature and are also from the authors upon request.

## Code availability

Open access software was used for the analysis.

## Author Contributions

Mahjani had full access to all the data in the study and takes responsibility for the integrity of the data and the accuracy of the data analysis.

*Study concept and design:* Akre, Batuure, Buxbaum, Grice, Gustavsson Mahjani, Janecka, Koskela, Mahjani

*Acquisition, analysis, or interpretation of data:* Akre, Buxbaum, Grice, Janecka, Koskela, Mahjani

*Drafting of the manuscript:* Batuure, Buxbaum, Grice, Gustavsson Mahjani, Janecka, Koskela, Mahjani

*Critical revision of the manuscript for important intellectual content:* All authors.

*Statistical analysis:* Mahjani

*Obtained funding:* Grice

*Study supervision:* Akre, Buxbaum, Grice, Koskela

---

Systematic Review and Meta-Analysis: Relationships Between Attention Deficit Hyperactivity Disorder and Daytime Urinary Symptoms in Children Supplements:

-Supplement 1: PRISMA checklist

-Supplement 2: Database search strategy

-Figure S1. Funnel plot

**Supplement 1:** PRISMA 2009 checklist.

| Section/topic | # | Checklist item | Reported on page # |
| --- | --- | --- | --- |
| **TITLE** |  |  |  |
| Title | 1 | Identify the report as a systematic review, meta-analysis, or both. | 1 |
| **ABSTRACT** |  |  |  |
| Structured summary | 2 | Provide a structured summary including, as applicable: background; objectives; data sources; study eligibility criteria, participants, and interventions; study appraisal and synthesis methods; results; limitations; conclusions and implications of key findings; systematic review registration number. | 2 |
| **INTRODUCTION** |  |  |  |
| Rationale | 3 | Describe the rationale for the review in the context of what is already known. | 3 |
| Objectives | 4 | Provide an explicit statement of questions being addressed with reference to participants, interventions, comparisons, outcomes, and study design (PICOS). | 3 |
| **METHODS** |  |  |  |
| Protocol and registration | 5 | Indicate if a review protocol exists, if and where it can be accessed (e.g., Web address), and, if available, provide registration information including registration number. | 5 |
| Eligibility criteria | 6 | Specify study characteristics (e.g., PICOS, length of follow-up) and report characteristics (e.g., years considered, language, publication status) used as criteria for eligibility, giving rationale. | 6 |
| Information sources | 7 | Describe all information sources (e.g., databases with dates of coverage, contact with study authors to identify additional studies) in the search and date last searched. | 5 |
| Search | 8 | Present full electronic search strategy for at least one database, including any limits used, such that it could be repeated. | 5<br>Supplement 2 |
| Study selection | 9 | State the process for selecting studies (i.e., screening, eligibility, included in systematic review, and, if applicable, included in the meta-analysis). | 5 |
| Data collection process | 10 | Describe method of data extraction from reports (e.g., piloted forms, independently, in duplicate) and any processes for obtaining and confirming data from investigators. | 6 |
| Data items | 11 | List and define all variables for which data were sought (e.g., PICOS, funding sources) and any assumptions and simplifications made. | 6 |
| Risk of bias in individual studies | 12 | Describe methods used for assessing risk of bias of individual studies (including specification of whether this was done at the study or outcome level), and how this information is to be used in any data synthesis. | 6 |
| Summary measures | 13 | State the principal summary measures (e.g., risk ratio, difference in means). | 6 |
| Synthesis of results | 14 | Describe the methods of handling data and combining results of studies, if done, including measures of consistency (e.g., $I^2$) for each meta-analysis. | 6 |
| Risk of bias across studies | 15 | Specify any assessment of risk of bias that may affect the cumulative evidence (e.g., publication bias, selective reporting within studies). | 6 |
| Additional analyses | 16 | Describe methods of additional analyses (e.g., sensitivity or subgroup analyses, meta-regression), if done, indicating which were pre-specified. | 6 |
| **RESULTS** |  |  |  |
| Study selection | 17 | Give numbers of studies screened, assessed for eligibility, and included in the review, with reasons for exclusions at each stage, ideally with a flow diagram. | 6, Figure 1 |
| Study characteristics | 18 | For each study, present characteristics for which data were extracted (e.g., study size, PICOS, follow-up period) and provide the citations. | 6-9<br>Table 1 |
| Risk of bias within studies | 19 | Present data on risk of bias of each study and, if available, any outcome level assessment (see item 12). | Figure 2 |
| Results of individual studies | 20 | For all outcomes considered (benefits or harms), present, for each study: (a) simple summary data for each intervention group (b) effect estimates and confidence intervals, ideally with a forest plot. | 9, Figure 2, |
| Synthesis of results | 21 | Present results of each meta-analysis done, including confidence intervals and measures of consistency. | Figure 2 |
| Risk of bias across studies | 22 | Present results of any assessment of risk of bias across studies (see Item 15). | 9, Figure 2,<br>Figure S1 |
| Additional analysis | 23 | Give results of additional analyses, if done (e.g., sensitivity or subgroup analyses, meta-regression [see Item 16]). |  |
| **DISCUSSION** |  |  |  |
| Summary of evidence | 24 | Summarize the main findings including the strength of evidence for each main outcome; consider their relevance to key groups (e.g., healthcare providers, users, and policy makers). | 9-10 |
| Limitations | 25 | Discuss limitations at study and outcome level (e.g., risk of bias), and at review-level (e.g., incomplete retrieval of identified research, reporting bias). | 10 |
| Conclusions | 26 | Provide a general interpretation of the results in the context of other evidence, and implications for future research. | 10-11 |
| **FUNDING** |  |  |  |
| Funding | 27 | Describe sources of funding for the systematic review and other support (e.g., supply of data); role of funders for the systematic review. | 3 |

From: Moher D, Liberati A, Tetzlaff J, Altman DG, The PRISMA Group (2009). Preferred Reporting Items for Systematic Reviews and Meta-Analyses: The PRISMA Statement. PLoS Med 6(7): e1000097. doi:10.1371/journal.pmed1000097

## Supplement 2: Database search strategy

**PubMed**

((urinary) OR (lower urinary tract) OR (overactive bladder) OR (bladder Pain Syndrome) OR (incontinence)) AND (attention deficit hyperactivity)) AND (urinary[Title] OR lower urinary tract[Title] OR overactive bladder[Title] OR bladder pain syndrome[Title] OR incontinence[Title] OR attention deficit hyperactivity[Title]) NOT rats[Title] NOT rat[Title] NOT dog[Title] NOT dogs[Title] NOT rabbits[Title] NOT rabbit[Title] NOT mice[Title]

Note: Similar strategies were used to search in PsycNet and Cochrane.

**Google Scholar**

We used Google Scholar to find articles that PubMed, PsycNet, and Cochrane searches might have missed. We performed 2 searches using the keywords below. For each search, we screened the first 100 results to find any relevant articles that PubMed, PsycNet, and Cochrane searches did not identify.

1. “attention deficit hyperactivity” “overactive bladder”

2. “attention deficit hyperactivity” “urinary”

**Figure S1.**
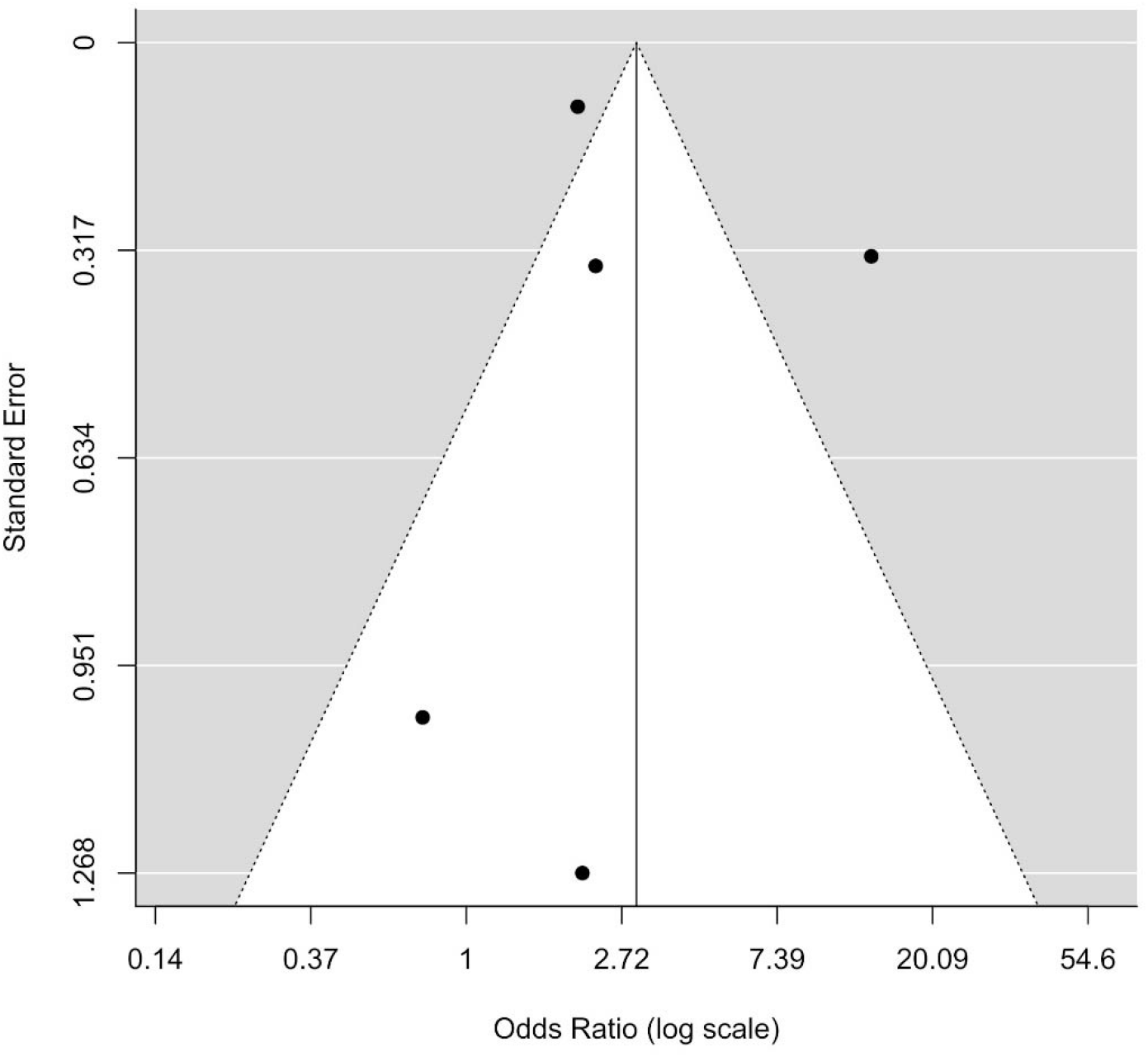
Funnel plot

